# Pandemic-EBT and grab-and-go school Meals: Costs, reach, and benefits of two approaches to keep children fed during school closures due to COVID-19

**DOI:** 10.1101/2022.04.06.22273512

**Authors:** Erica L. Kenney, Lina Pinero Walkinshaw, Ye Shen, Sheila E. Fleischhacker, Jessica Jones-Smith, Sara N. Bleich, James W. Krieger

**Affiliations:** Department of Nutrition, Harvard T.H. Chan School of Public Health, 665 Huntington Avenue, Boston, MA, 02115; University of Washington, Department of Health Systems and Population Health; Center for Health Decision Science, Harvard T.H. Chan School of Public Health; Georgetown University Law Center, Washington, DC; University of Washington, Department of Epidemiology; Department of Health Policy and Management, Harvard T.H. Chan School of Public Health

## Abstract

**Importance:** School meals improve nutrition and health for millions of U.S. children. School closures due to the COVID-19 pandemic disrupted children’s access to school meals. Two policy approaches were activated to replace missed meals for children from low-income families. The Pandemic Electronic Benefit Transfer (P-EBT) program provided the cash value of missed meals directly to families on debit-like cards to use for making food purchases. The grab-and-go meals program offered prepared meals from school kitchens at community distribution points. The effectiveness of these programs at reaching those who needed them and their costs were unknown.

**Objective:** To determine how many eligible children were reached by P-EBT and grab-and-go meals, how many meals or benefits were received, and how much each program cost to implement.

**Design:** Cross-sectional study, Spring 2020.

**Setting:** National.

**Participants:** All children <19 years old and children age 6-18 eligible to receive free or reduced price meals (FRPM).

**Exposure(s):** Receipt of P-EBT or grab-and-go school meals.

**Main Outcome(s) and Measure(s):** Percentage of children reached by P-EBT and grab-and-go school meals; average benefit received per recipient; and average cost, including implementation costs and time costs to families, per meal distributed.

**Results:** Grab-and-go school meals reached about 10.5 million children (17% of all US children), most of whom were FRPM-eligible students. Among FRPM-eligible students only, grab-and-go meals reached 27%, compared to 89% reached by P-EBT. Among those receiving benefits, the average monthly benefit was larger for grab-and-go school meals ($148) relative to P-EBT ($110). P-EBT had lower costs per meal delivered - $6.51 - compared to $8.20 for grab-and-go school meals. P-EBT had lower public sector implementation costs but higher uncompensated time costs to families (e.g., preparation time for meals) compared to grab-and-go school meals.

**Conclusions and Relevance:** Both programs supported children’s access to food when schools were closed and in complementary ways. P-EBT is an efficient and effective policy option to support food access for eligible children when school is out.

**KEY POINTS:** *Question:* What were the operating costs, costs and benefits to families, and proportion of eligible children who received benefits of two programs aimed at replacing school meals missed when schools were closed due to COVID-19?

*Findings:* In this cross sectional analysis, we found that the Pandemic-Electronic Benefit Transfer program, in which state agencies sent debit cards loaded with the cash value of missed school meals directly to families, reached nearly all low income students (89%) and cost relatively little per meal provided. In comparison, grab-and-go school meals, in which school food service departments provided prepared meals for offsite consumption, reached 27% of low income children and was associated with larger per meal costs.

*Meaning:* During times when children cannot access school meals, state and federal agencies should support cost-efficient programs for schools to distribute prepared meals and activate programs like P-EBT to efficiently reach eligible children.

## INTRODUCTION

Ensuring children have adequate nutrition—continuous access to enough healthy food to promote healthy development—is a critical public health challenge.(1)(2,3) Inequitable access to healthy food produces disparities in diet quality that lead to racial and socioeconomic health inequities from early ages.(4–6) Children from low-income households are at heightened risk for experiencing food insecurity and inadequate nutrition (7), bear a disproportionate burden of childhood obesity (8,9) and thus experience multiple nutrition-related threats to their health.(10)

The National School Lunch and Breakfast Programs (NSLP/SBP), administered by the United States Department of Agriculture (USDA), are powerful tools for improving children’s nutrition and reducing health inequities. They significantly reduce food insecurity,(11,12) improve diet quality, and reduce obesity risk for low income children.(13–15) Before the COVID-19 pandemic, over 30 million children received NSLP meals each year, 22 million of whom were from households with incomes ≤185% of the Federal poverty level, thus qualifying them for free or reduced price meals (FRPM). (16)

However, school closures due to the COVID-19 pandemic in spring 2020 disrupted access to school meals for millions of US children, sharply increasing the risk of food insecurity for children depending on this food source. In response, Congress authorized the USDA to implement two approaches: 1) grab-and-go school meals, in which school food authorities switched from preparing meals for students to eat inside schools to distributing prepared meals for offsite consumption via community distribution sites or mobile delivery systems, and 2) the Pandemic-Electronic Benefit Transfer (P-EBT) program, in which states distributed the cash value of missed school meals to parents of FRPM-eligible children on a debit-like card so they could purchase groceries from food retailers, similar to EBT cards used in the Supplemental Nutrition Assistance Program (SNAP). Preliminary reports suggest both programs helped children replace food from missed school meals and alleviated household food insecurity.(17,18) However, it is not clear how effective the programs were at reaching and distributing benefits to eligible children during school closures, or how much they cost.

This study’s aim was to estimate the effectiveness in reaching FRPM-eligible children, program implementation and family costs, benefits received by participating children, and cost per meal distributed for the two programs during the spring of 2020, a period when nearly all schools across the nation were closed.(19)

## METHODS

### Study design and population

This cross-sectional analysis used a framework developed by Dietz and Gortmaker(20) to evaluate the costs, population reach, and benefits distributed of P-EBT and grab-and-go school meals. Our primary analysis examined the extent to which these programs reached children aged 6-18 living in the United States who were eligible for FRPM, as this was the population most affected by school closures. This population included both children who were eligible because their families had incomes ≤185% of FPL as well as children attending school districts that participated in the Community Eligibility Program (in which districts with large proportions of low income children offer free meals to all children regardless of income). Because schools provided grab-and-go meals to all children <19 years old (regardless of income or CEP participation), we also evaluated how effectively this meal program reached this broader population. We leveraged multiple government and administrative datasets with national- and state-level data on program participation, costs, and benefits for the spring of 2020 (Table 1). Analyses included all states except for South Dakota and Wyoming, which were excluded due to insufficient data.

**Table 1.** Data sources for estimating program reach, benefits, and implementation costs for grab-and-go school meals and the Pandemic Electronic Benefit Transfer (P-EBT) programs, Spring 2020.

| Source | Data Description |
| --- | --- |
| **REACH** |  |
| U.S. Census Household Pulse Survey(24) | Number and proportion of free and reduced-price meal (FRPM) eligible families who report picking up a free school meal. |
| USDA P-EBT 2019-2020 SY State Plans(22) | Total number of P-EBT/FRPM-eligible children per state (for some states) |
| American Community Survey(23) | Number of children age 0-19 per state |
| Center on Budget and Policy Priorities and Food Research & Action Center, “Pandemic-EBT Implementation Documentation Project”(21) | Surveys of total number of P-EBT/FRPM-eligible children; number of times P-EBT benefits were issued in each state |
| State government websites and press releases | Number of children receiving P-EBT |
| USDA P-EBT Distribution Data(25) | Number of children receiving P-EBT |
| **BENEFITS** |  |
| USDA Child Nutrition Tables (School Meal Distribution and Reimbursement Data for NSLP, NSBP, SSO, SFSP) | Total meals distributed in each state for April and May of 2020 |
| USDA P-EBT Distribution Data(25) | Total dollar amounts disbursed per state per month for March – June 2020 |
| USDA P-EBT 2019-2020 SY State Plans(22) | Planned/budgeted dollar amounts to be disbursed for P-EBT for March- June 2020 |

| **COSTS** |  |
| --- | --- |
| USDA P-EBT 2019-2020 SY Approved State Plans(22) | State-requested P-EBT administrative funding for the 2020-2021 academic school year. |
| USFA School Meal Cost Survey(29) | District-level cost associated with delivering school meals (including administrative, operating, and food costs) for 7 of the largest US school food authorities. |
| UW School Meal Cost Survey (primary data) | District-level cost associated with delivering school meals (including administrative, operating, and food costs) for a convenience sample of 17 districts nationally. |
| Davis & You (2010): Family cooking time and meal prep cost estimates(31) | Estimated cost to FRPM-eligible family (including time and wage) to prepare home meals. |
| Voulgaris et al (2017) Travel to school time and mileage to school estimates(30) | Estimated travel cost for a family to reach a school site to pick up a school meal to go (including time and mileage costs, taking into account drivers and bus-riders.) |

### Program reach

Program reach is defined as the proportion of children eligible for each program who receive benefits. The eligible population for P-EBT was FRPM-eligible students. The eligible population for grab-and-go meals was all children under 19 years old, and we also focused on the subset of this population that was FRPM-eligible students. The number of FRPM-eligible students in each state was obtained from state applications to USDA for P-EBT funding combined with survey data.(21) (22) The number of all children under 19 years old was obtained from the American Community Survey.(23)

To estimate how many children both nationally and in each state benefitted from grab-and-go school meals in spring of 2020, we used data from the Census Household Pulse Survey(24), which collected weekly nationally representative data on household composition, income, and whether or not a household had received free meals from schools; we stratified these estimates by state. To estimate how many children received P-EBT benefits, we triangulated data from: (a) state government websites or data released via a Freedom of Information Act request (available from 22 states); (b) states’ estimates to USDA of the amount of cash they planned to disburse for spring 2020 and the total number of days benefits were issued; (22) and (c) USDA-released data on monthly estimates of benefit receipt.(25)

More details can be found in the **Appendix**.

### Program benefit*s*

Program benefits for participating children were estimated in two ways: as the monthly cash value of the benefits and the number of meals or meal-equivalents received. The number of meals provided nationally and per state by grab-and-go school meals programs for April and May of 2020, by FRPM-eligibility, was obtained from administrative data provided by USDA. The cash value of benefits provided per state for P-EBT was estimated by triangulating data from states’ P-EBT applications to USDA(22) and administrative data from USDA on benefit disbursal for March-June 2020(25). We converted the meals distributed through grab-and-go to their cash value using their USDA reimbursement value for 2020 ($5.85 for two meals per day for continental U.S.). Similarly, we converted P-EBT cash benefits to meal-equivalents by dividing the total cash benefits distributed by the average cash benefit distributed per day ($5.70 for continental U.S.), then multiplying by two, since two meals per day were meant to be covered. If the estimate for the average number of meals distributed per child per month for grab-and-go meals for a state was greater than 60 (the maximum possible number, given two meals per a 30 day month), we capped this measure at 60 meals per month (done for 14 states). Similarly, for P-EBT, if a state estimate was greater than 40 meals (the maximum possible meal-equivalents), we capped the value at 40 (done for 28 states). More details on these methods can be found in the **Appendix**.

### Cost per meal delivered

#### Program costs: Grab-and-go meals

We followed standard guidelines for identifying, measuring, and valuing the resources required for program costs.(26) (27,28) We combined data from an existing study of school food costs(29) with data we collected from a survey of a convenience sample of school food service directors from 17 districts, resulting in a total sample of 24 school districts across nine states. Data included information on labor (including fringe benefits), food (including food waste), and operating costs (such as personal protective equipment, transportation, and takeout containers) associated with grab-and-go meal implementation. The surveys also captured the number of meals served in each district, allowing us to standardize costs per-meal. We additionally accounted for uncompensated costs to families, namely, the average time spent and travel costs expended for families to pick up meals, using previously published estimates of the travel time for students to their local schools(30) and the value of adult caregivers’ time in FRPM-eligible households (31). These costs were then weighted to estimate average state- and national-level costs. More details on the calculation of grab-and-go meal costs can be found in the **Appendix**.

#### Program costs: P-EBT

To estimate the costs of P-EBT, we used each state’s P-EBT application to the USDA,(22) which included estimates of state-specific labor costs for identifying and contacting eligible participants, distributing benefits, and monitoring the program, as well as the cash value of the P-EBT benefits disbursed and equipment costs (processing and mailing of EBT cards). We then summed costs across all states to calculate national program costs. For uncompensated costs to families, we accounted for time spent by family members to prepare meals with foods purchased through P-EBT using existing estimates of preparation time for breakfasts and lunches in low income households(32) and the same valuation of FRPM-eligible caregivers’ time as above.(31)

#### Cost per meal

To calculate the cost per meal delivered for each program from a societal perspective, we divided the national monthly cost of meals distributed by the national monthly number of meals or meal-equivalents delivered per month. A secondary analysis separately calculated the cost per meal provided for public agencies and families.

Analyses were conducted using Microsoft Excel and StataMP 14.(33)

## RESULTS

In spring 2020, approximately 8 million of the 30 million FRPM-eligible in the US were reached by the grab-and-go school meals program (27%). The program reached an additional 2.5 million children who were not eligible for FRPM. The reach of grab-and-go meals for FRPM-eligible children varied substantially by state, ranging from 14% to 54% (**Table 2 and Figure 1**). Meanwhile, P-EBT reached 26.9 million FRPM-eligible children nationally (89%), with the percentage reached also varying substantially across states, from 51 to 100%.

**Table 2.**
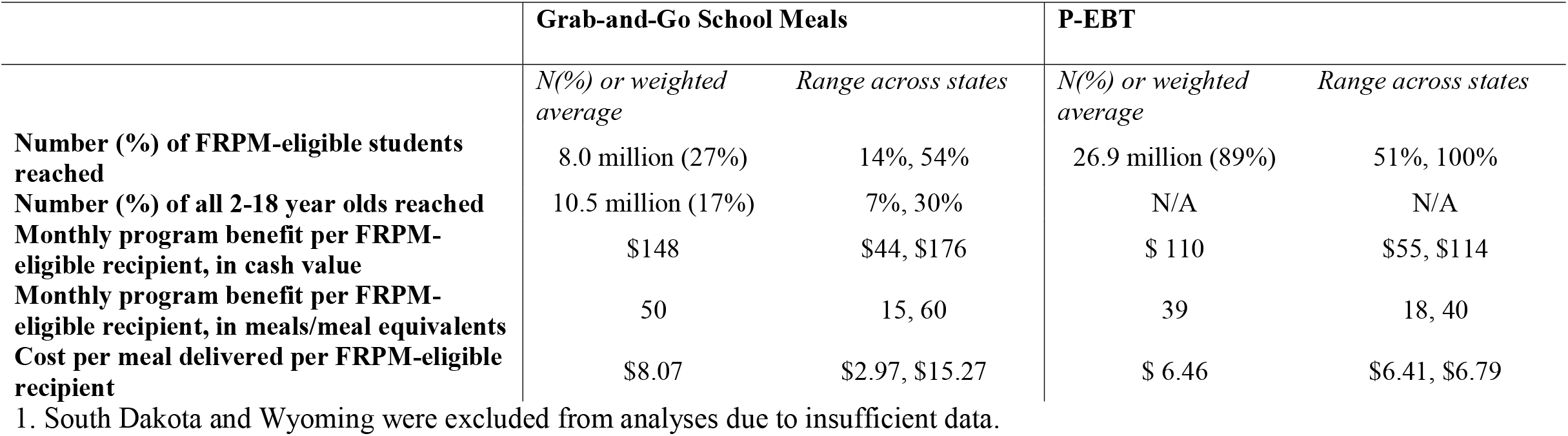
National^1^ reach, benefits, and implementation costs per month for grab-and-go school meals and the Pandemic Electronic Benefit Transfer (P-EBT) programs, Spring 2020.

|  | **Grab-and-Go School Meals** |  | **P-EBT** |  |
| --- | --- | --- | --- | --- |
|  | *N(%) or weighted average* | *Range across states* | *N(%) or weighted average* | *Range across states* |
| **Number (%) of FRPM-eligible students reached** | 8.0 million (27%) | 14%, 54% | 26.9 million (89%) | 51%, 100% |
| **Number (%) of all 2-18 year olds reached** | 10.5 million (17%) | 7%, 30% | N/A | N/A |
| **Monthly program benefit per FRPM-eligible recipient, in cash value** | \$148 | \$44, \$176 | \$ 110 | \$55, \$114 |
| **Monthly program benefit per FRPM-eligible recipient, in meals/meal equivalents** | 50 | 15, 60 | 39 | 18, 40 |
| **Cost per meal delivered per FRPM-eligible recipient** | \$8.07 | \$2.97, \$15.27 | \$ 6.46 | \$6.41, \$6.79 |

1. South Dakota and Wyoming were excluded from analyses due to insufficient data.

**Figure 1.**
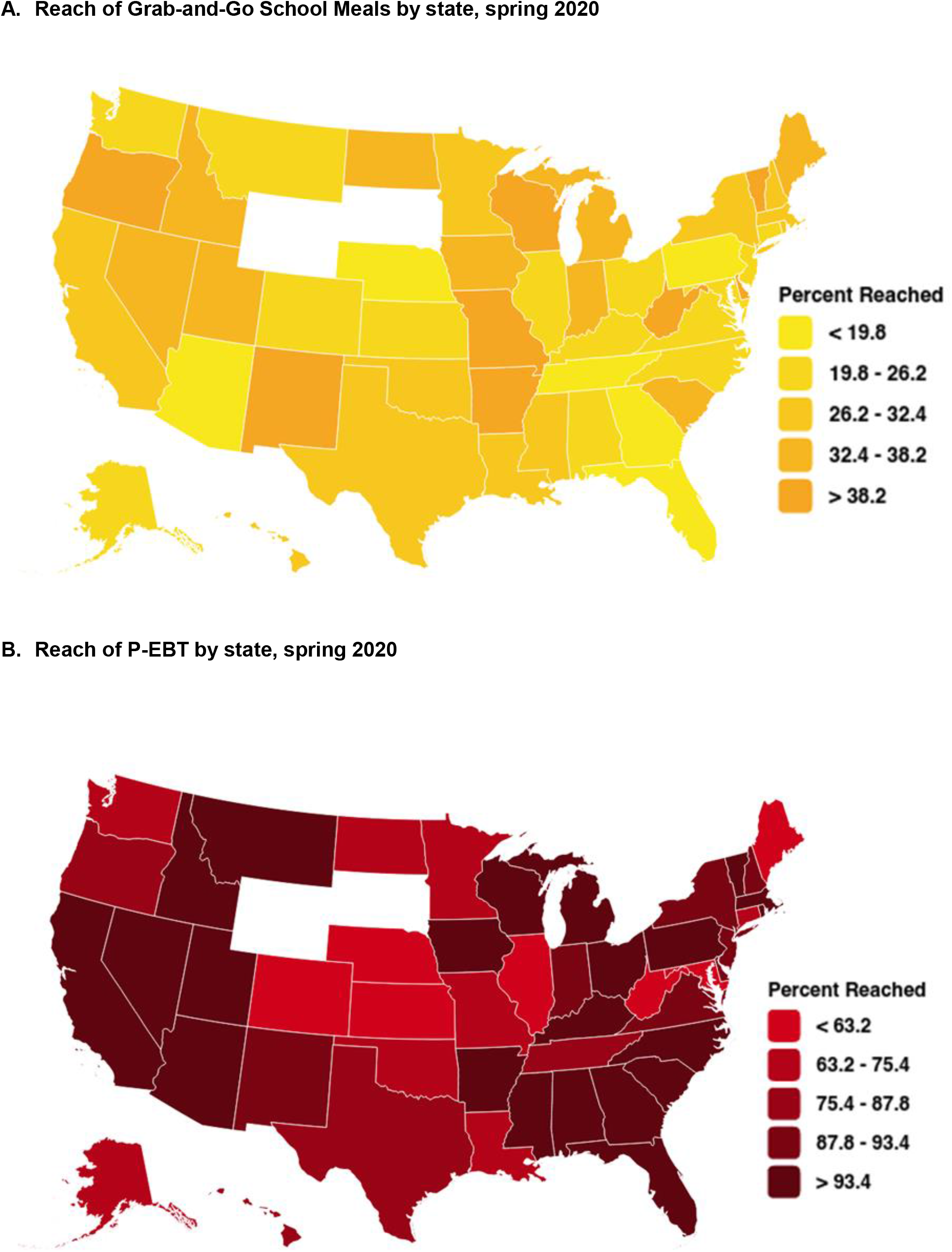
Percentage of free-and-reduced price meal eligible school students participating in (A) grab- and-go school meals and (B) Pandemic-Electronic Benefit Transfer (P-EBT) programs by state, United States, Spring 2020.

The grab-and-go school meals program distributed an average of 429 million meals per month with an estimated value of $1.2 billion in spring 2020. This translated to an average of 50 meals distributed per month per FRPM-eligible child (range across states of 15 – 60), with a retail cash value of approximately $148 per month per child (range across states: $44 - $176).

States issued an average of $3.2 billion in monthly cash benefits, equivalent to 1.1 billion meals, through P-EBT in spring 2020 (although not all these benefits were actually received by families during this time period – in many states, benefits did not actually get to families until as late as August or September of 2020 due to P-EBT implementation delays). This translated to an estimated $110 in cash-value benefits per month per child receiving P-EBT benefits (range across states: $55 - $114), or approximately 39 meals that could potentially have been purchased with P-EBT benefits per month per child (range across states: 18-40).

The total national average weighted cost per meal provided was $8.07 for grab-and-go school meals and $6.46 for P-EBT. The uncompensated cost for families for grab-and-go meals ($1.00 per meal) was lower than for P-EBT ($3.56 per meal). The public agency cost component was lower for P-EBT ($2.90 per meal) relative to grab-and-go school meals ($7.07).

## DISCUSSION

School meal programs play an essential role in ensuring that millions of US children, particularly those living in households near or in poverty, have access to meals that promote healthy growth and development.(34) For most US children, a school meal is the most nutritious meal of their day.(35) The best approaches to ensuring access to nutritious meals when schools are closed, whether during emergencies or routine closures such as summer vacation, are thus an important policy question. This study suggests that P-EBT can reach the vast majority of eligible children at relatively low cost to the government, while a meal distribution model such as grab-and-go school meals can also ensure families directly receive meals and reach children beyond those who are FRPM-eligible. Our results, as well as those of other researchers, suggest that disaster preparedness plans that help school meal programs safely set up community distribution sites (29,36–39) and establish infrastructure so that P-EBT can be rapidly deployed(39) are needed to prevent future disruption of food access for children. Our P-EBT findings also suggest that there may be a benefit to scaling up the USDA’s Summer-EBT program, which is currently a pilot program,(40) to a national scale.

Although P-EBT was implemented at varying paces across the states,(21) with many families not receiving the benefits allotted to them for spring 2020 until later in the year, our analysis suggests that during a time of uniform school closures P-EBT was effective at reaching most FRPM-eligible children. P-EBT provided benefits to 89% of eligible children, compared to 27% for grab-and-go school meals. Grab-and-go school meals may have reached fewer families because they had to travel to a distribution site at fixed times to obtain meals. Travel was often a challenge for them(39), despite substantial efforts by school food authorities to make meal acquisition as easy as possible.(29,37) In contrast, the effort required for households to receive P-EBT was relatively low, in that cards were automatically mailed to families of children known to be eligible and could be used to buy food at any store accepting EBT. It is worth noting that a recent report suggested that the high P-EBT coverage seen when schools were uniformly closed may be difficult to maintain during periods when the extent of school closures varies. In the latter context, it is more challenging to determine number of missed meals for each child, leading some states to forgo reapplying to USDA to continue the program.(41)

In this study, the per-meal cost of P-EBT ($6.46) was less than that of grab-and-go school meals ($8.07). P-EBT’s lower costs appear related to the fewer resources required for implementation. In contrast, grab-and-go school meals required more resources, particularly labor and equipment, due to the need to source, prepare, package, distribute, and seek reimbursement for meals.(42) Prior research has shown that delivery of grab-and-go school meals is more costly compared to providing regular school meals on-site when schools are open.(29).

While our analyses suggest P-EBT reached more FRPM-eligible children at a lower cost per meal, P-EBT should not be the sole response to address children’s meal access when schools are closed. Given the initial delays in implementation of P-EBT as well as potential difficulties in implementing the program when schools are not uniformly closed, P-EBT may not, in its current configuration, quickly reach all students. The program may need reconfiguration to simplify determination of missed meals and streamline benefit distribution.

Grab-and-go school meals were also an important safety net for families cut off from school meals but not able to access P-EBT, including the roughly 3.5 million households with food-insecure children that are not income-eligible for FRPM,(7) families that experienced sudden loss of income due to the pandemic that made them eligible for P-EBT but who might not have been captured by administrative data as being eligible, and those without stable housing and accurate mailing addresses to allow delivery of P-EBT cards. Grab-and-go school meals also may have reduced the time cost of preparing meals for children, which can be quite high for low-income caregivers.(31) School employees have noted that grab-and-go school meals programs allowed school staff to maintain contact with families who would otherwise have been isolated and to distribute home-learning resources. (29) Lastly, the grab-and-go school meals had to meet at least basic USDA nutritional standards, while P-EBT could be used to purchase any type of food. Thus, while P-EBT gave parents more choice, the foods purchased with P-EBT benefits may not have been as nutritionally sound as grab-and-go school meals.

Strengths of this study include the use of detailed administrative data, the triangulation of data gathered from multiple sources to maximize data validity, and collection of detailed cost data for grab-and-go school meals. However, our study has several limitations related to the available data sources. First, some states’ P-EBT administrative data did not separate distribution of P-EBT and SNAP benefits, which made it impossible to accurately estimate the number of unique P-EBT recipients and the benefits they received using a single data source for those states, making it necessary to triangulate several data sources. In some states, estimates of benefits provided by both P-EBT and grab-and-go meals exceeded the maximum permitted by USDA, leading us to cap estimated benefits distributed per person if they exceeded this threshold. High estimates may have resulted from the previously mentioned limitations of P-EBT data, errors in states’ reports of distributed grab-and-go meals, or errors in estimates of FRPM-eligible students. Census Pulse Household Survey data, which we used to estimate the number of children receiving grab-and-go school meals, assessed receipt at the household, not child, level. Finally, we were unable to locate a source of data for federal program administrative costs (i.e., at USDA). However, these costs were likely similar for the two programs and small compared to the larger cost contributions attributable to the benefits themselves and labor costs.

Our study examined the start-up period of both programs. Additional analysis is needed to assess program reach when school closures are more inconsistent and whether costs might decrease once programs are mature and stabilized. Indeed, the range in estimated costs across states suggests that some states may have developed particularly cost-efficient approaches for program implementation. Identifying these best practices for minimizing program costs could inform future program implementation.

Lastly, we were unable to identify data to assess the impact of the programs on children’s food and nutrition security. It would be useful to understand the extent to which P-EBT benefits were redeemed, whether they were used to buy food for household children, and the nutritional quality of the foods purchased with them. Similarly, future research should evaluate the degree to which grab-and-go school meals were consumed, who consumed them, and their effects on diet quality.

## CONCLUSIONS

School closures in response to the COVID-19 pandemic created a challenge for children and families who depend on school meals. The combination of P-EBT and grab-and-go school meals offers a two-pronged strategy to facilitate food access when schools are closed, whether during emergencies or routine breaks. The ability of P-EBT to reach nearly all eligible students from low income households could prevent gaps in food access due to school closures, while grab-and-go school meals could reach households that are not eligible for EBT benefits and provide meals that meet USDA nutrition standards. This study, with its findings on the reach, costs, and benefits of each approach, suggests that future development of the programs could consider 1) investigating strategies for cost containment for grab-and-go school meals; 2) expanding P-EBT to cover 60 meals per month instead of 40, in order to match the grab-and-go school meals benefit level; 3) exploring the use of P-EBT during all times when schools are closed for lengthy periods (e.g. summer vacations), not just emergencies; and 4) optimizing the nutritional quality of the foods provided.

## Data Availability

All data produced in the present study are available upon request to the authors

## Acknowledgments

The authors wish to thank members of the Nutrition and Obesity Policy Research and Evaluation Network (NOPREN) Food Security Work Group as well as the HER/NOPREN COVID-19 Food and Nutrition (COVID-19 F&N) Work Group for their insights and feedback on earlier presentations of these analyses. The authors also wish to thank Kyla Tucker, MPH, for assistance with data collection.

## Disclaimers

The views expressed in this manuscript by Drs. Bleich and Fleischhacker, who each transitioned to the federal government during the later stages of this study, are solely the personal views of those authors.

