## Appendix for "Pandemic-EBT and grab-and-go school Meals: Costs, reach, and benefits of two approaches to keep children fed during school closures due to COVID-19"

**P-EBT and Grab-and-go Meals Cost Effectiveness Analysis: APPENDIX**

*Contents*:

1) Formulas for calculating metrics
2) Methods for estimating proportion of families collecting grab-and-go meals
3) Decision process for selecting sources for P-EBT reach and benefits
4) Calculation of grab-and-go school meals costs
5) Calculation of P-EBT costs

### A1. Formulas for calculating metrics

| **Monthly program reach** | |
| --- | --- |
|  | **FRPM-eligible** |
| **School Meal** | (proportion of FRPM students receiving school MTG, Census Pulse)*(N of FRPM eligible students, FRAC estimate + ACS 2-5 not in public school FRPM-eligible) |
| **School Meal** | (proportion of FRPM students receiving school MTG, Census Pulse)*(N of FRPM eligible students, FRAC estimate) |
| **PEBT** | (N of students receiving PEBT, USDA data) / (N of eligible students, FRAC estimate) |
| **Monthly individual program benefit for children receiving benefit, in cash value** | |
|  | **FRPM-eligible** |
| **School Meal** | (((N FRP meals distributed in [month])/2)*5.85) /(proportion FRP students received meals, Census Pulse * N of FRPM eligible students, FRAC) |
| **PEBT** | (total cash benefits distributed for P-EBT [month], USDA data) / (N students receiving P-EBT [month], USDA data) |
| **Monthly individual program benefit for eligible children, in cash value** | |
|  | **FRPM-eligible** |
| **School Meal** | (((N FRP meals distributed in [month])/2)*5.85) / N FRP students eligible for meals, FRAC estimate |
| **PEBT** | (total cash benefit distributed for PEBT [month], USDA data) / (N students eligible for PEBT [month], USDA data) |
| **Monthly program benefit per student receiving meal, in meals** | |
|  | **FRPM-eligible** |
| **School Meal** | (N FRP meals distributed in [month], USDA data) / (proportion FRP students receiving meals, Pulse * number of FRP eligible students, FRAC) |
| **PEBT** | ((total cash benefits distributed in [month], USDA data)/(5.7/2)) / (N students receiving benefits, USDA data) |
| **Monthly program benefit per eligible student, in meals** | |
|  | **FRPM-eligible** |
| **School Meal** | (N FRP meals distributed in [month], USDA data) / N FRP students eligible for meals, FRAC estimate |
| **PEBT** | ((total cash benefits distributed in [month], USDA data)/(5.7/2)) / (N students eligible for PEBT [month], USDA data) |
| **Cost per meal delivered** | |
|  | **FRPM-eligible** |
| **School Meal** | (monthly program costs, including all framework elements) / N monthly ALL meals distributed |
| **PEBT** | (monthly program costs, including all framework elements) / ((total cash benefits distributed in [month], USDA data)/(5.7/2)) |
| **Monthly cost per benefiting child** | |
|  | **FRPM-eligible** |
| **School Meal** | (total monthly program costs, including all framework elements) / (proportion ALL students receiving meals, Pulse* N all students, NCES) |
| **PEBT** | (total monthly program costs, including all framework elements) / (N all students receiving PEBT benefits, USDA) |

### A2. Methods for estimating reach of grab-and-go meals

To estimate the proportion of children receiving benefits through grab-and-go school meals, we used data from the Census Household Pulse Survey,(25) which collected weekly repeated cross-sectional estimates of different indicators of population well-being and economic status at the household level throughout the COVID-19 pandemic, beginning April 23, 2020. We used the following questions from the survey to develop our estimates:

| **Variables used from U.S. Census Household Pulse Survey (spring 2020):** | |
| --- | --- |
| Variable name | Question and response options |
| THHLD_NUMPER | Total number of people in household |
| THHLD_NUMKID | Total number of people under 18 years old in household |
| FREEFOOD | During the last 7 days, did you or anyone in your household get free groceries or a free meal? (Yes/No) |
| WHEREFREE | (If Yes): Where did you get free groceries or free meals? Choose all that apply.  - Free meals through the school or other programs aimed at children  -Food pantry or food bank  - Home-delivered meal service like Meals on Wheels  -Church, synagogue, temple, mosque, or other religious organization  -Shelter or soup kitchen  -Other community program  -Family , friends, or neighbors |
| ENROLL1 | "At any time during February 2020, were any children in this household enrolled in a public  school, enrolled in a private school, or educated in a homeschool setting in  Kindergarten through 12th grade or grade equivalent? Select all that apply. - Yes, enrolled in a public or private school" |
| INCOME | In 2019 what was your total household income before taxes? Select only one answer:  "1) Less than $25,000  2) $25,000 - $34,999  3) $35,000 - $49,999  4) $50,000 - $74,999  5) $75,000 - $99,999  6) $100,000 - $149,999  7) $150,000 - $199,999  8) $200,000 and above |

To classify households as either FRPM-eligible or non-eligible, we used the income and household size variables to estimate the household’s income-to-poverty ratio using guidance for calculating poverty thresholds from the U.S. Census (<https://www.census.gov/data/tables/time-series/demo/income-poverty/historical-poverty-thresholds.html>). We then set all households with a ratio ≤1.85 and reporting that they had a student attending school as eligible for FRPM.

To classify households as receiving grab-and-go meals or not, we coded all families that selected first that they had received free groceries or a free meal in the last 7 days and then selected that they had gotten the meals through the school or other programs aimed at children as having receive grab-and-go meals. While there may be some misclassification in this variable due to the phrase “other programs aimed at children,” this is likely to be quite small, given that the listed options for other named sources were comprehensive and likely to cover any non-school programs.

We then calculated a) the proportion of FRPM-eligible students receiving grab-and-go school meals and b) the proportion of all children <19 years old receiving grab-and-go meals. For the first, we summed the reported number of children in the households reporting receipt of school meals among FRPM-eligible households and then divided by the sum of children in all FRPM-eligible households. For the second, we summed the reported number of children in all households reporting receipt of school meals (regardless of FRPM-eligibility) and divided by the sum of the number of children in all households with children.

We calculated these metrics for six weeks, from April 23 – June 9^th^, stratified by state. Because reach was variable across weeks (possibly due to real differences in reach or initial difficulty getting the program running, but also possibly due to imprecision in state-specific estimates), we selected the highest reach estimate for each state among these weekly estimates to represent spring 2020 reach overall.

### A3. Decision process for selecting P-EBT reach data source

**SOURCES FOR ESTIMATING THE NUMBER OF BENEFITTING CHILDREN PER STATE, P-EBT, IN ORDER OF PREFERENCE (WITH SOME EXCEPTIONS)**

1) State press releases or state-released data from FOIAs submitted by investigators
2) Taking the total amount of cash benefits planned to be disbursed from March-Sept 2020 and dividing by the product of the maximum daily benefit amount times the number of days the state was issuing benefits, using states’ initial P-EBT plans submitted to and approved by USDA as the estimate of cash benefits
3) Summing the “people” estimates from USDA’s data release on P-EBT across March-Sept 2020, then dividing by the number of issuances for that state
4) Taking the total amount of cash benefits disbursed from March-Sept 2020 and dividing by the product of the maximum daily benefit amount times the number of days the state was issuing benefits, using data from the USDA data release (note: this is secondary to option 2 because we learned that many states had mixed in SNAP benefits in P-EBT data. However, there are a few instances where the planned estimates from option 2 are much too high and these option 4 estimates were more reasonable).
5) After reviewing monthly reimbursement data, identifying any patterns where it appeared that a single month’s number of “people” getting benefits might actually represent the entire population getting benefits, i.e. the states reported serving the same number of people each month

**DECISION FLOW FOR SELECTING THE MOST RELIABLE SOURCE**

Do we have an estimate from a state-specific agency press release or website announcing the number of children who received benefits for spring 2020?

🡪 if YES, then use this estimate (Option 1 above)

🡪 If NO, then:

- When summing the amount disbursed (estimated from what states proposed in USDA plans) from March-Sept and dividing by the product of the maximum daily benefit amount times the number of days the state was issuing benefits, do we have a reach estimate <100%?
  - If YES, then use this estimate (Option 2 above)
  - If NO, then:
- Is there an estimate when summing the “People” estimates from March-Sept 2020 from the USDA data release and dividing by estimates of number of issuances that is less than 100%?
  - IF YES, then use this estimate (Option 3 above)
  - IF NO, then consider:
    - Is the percent reached calculated from the amount disbursed at least less than <110%?
      - IF YES, then use this same estimate, truncating to 100%
      - IF NO, then consider:
        - Do we have an estimate from summing the amount disbursed from the USDA that is <100%?

If YES, then use this estimate (Option 4 above)

If NO, then consider:

Did we identify, in examining monthly reimbursement data, any patterns where a single month’s number of “people” getting benefits plausibly represents the population getting benefits?

If YES, then use this estimate (Option 5 above)

If NO, then EXCLUDE and STOP

### A4. Calculation of grab-and-go school meals costs

***Costs to school food authorities***

To estimate the costs to school food authorities to operate grab-and-go school meals, we used data collected in a prior study of large urban school food districts in the country (Kenney et al 2021), which included cost data for 7 districts, combined with a survey we administered to a convenience sample of school food service directors from 17 districts across 5 states. The data collected included:

- Number of students enrolled in the district

- Number of lunches served per month during school closures in 2020

- Number of breakfasts served per month during school closures in 2020

- Whether or not the district was charged with also serving meals to adults in need

- Total monthly labor costs including wages, overtime, hazard pay, overhead and benefits during school closures in 2020

- Breakfast food costs per month during school closures in 2020

- Lunch food costs per month during school closures in 2020

- Recurring equipment and supply costs, such as food packaging materials (e.g., carryout containers, bags, disposable silverware), staff personal protective equipment (e.g., face masks, face shields, gloves, sanitizing and cleaning supplies), on-site communication materials (e.g., signage, floor stickers), mailers, fliers, or other communication materials, and any other (per month during school closures in 2020)

- One-time equipment and supply costs, such as freezers, folding tables, or carts that were purchased to set up grab-and-go meal distribution (per month during school closures in 2020)

- Whether or not the district delivered meals or distributed via stationary sites

- (If delivery): Driver wages, benefits, and overtime if not already included above in ‘labor costs.’

- (If delivery): Costs associated with renting or operating delivery vehicles.

- (If delivery): Fuel expenses, if not included in the vehicle expenses above.

- Other –any other costs associated with meal delivery

From these data, we then calculated the total school food authority operating cost for each surveyed district per month during COVID-19 school closures, and divided this by the sum of the reported lunches and breakfasts served per month during COVID-19 school closures to calculate a per-meal cost. To calculate a national weighted average cost per meal, we first calculated cost per meal for each district providing data. We then created an average cost per meal within each of the nine participating states, weighting each district’s cost per meal by the total number of students in that district. Lastly, we calculated the average cost per meal for school food authorities across all nine states, weighted by the total number of FRPM-eligible students in that state. Treating these nine states as a sample of the U.S. states, we thus used this weighted average cost across the nine sampled states as the national weighted cost per meal.

***Time and travel costs for families (uncompensated):*** We calculated the time spent and travel costs expended on average for families to pick up grab-and-go school meals. We estimated that the average distance from a child’s home to their school was 4.8 miles; that this trip lasted 18 minutes on average(31); and that families made four trips per month (assuming a weekly pickup of meals). We used the 2020 federal mileage reimbursement rate to estimate the cost of a car trip and valued adult caregivers’ time in FRPM-eligible households using a market substitute based estimate developed by Davis & You (2010)(32) and inflating to 2020 dollars. We then weighted this estimate by the proportion of school districts that used a parent-pickup model as opposed to a delivery model using unpublished data collected via survey by Julia McCarthy and colleagues at the Laurie M. Tisch Center for Food, Education & Policy.

### A5. Calculation of P-EBT costs

**Costs for state governments:** To estimate the costs of P-EBT, we used each state’s P-EBT application to the USDA,(24) which included each state’s own estimates of their state agency-specific labor costs for identifying and contacting eligible participants, distributing benefits, and monitoring the program. The cash value of the P-EBT benefits disbursed and equipment costs (processing and mailing of EBT cards) were also included in the estimates from the state applications.

**Time costs for families (uncompensated):** We accounted for time spent by family members to prepare meals with foods and beverage purchased through P-EBT by using the same valuation of FRPM-eligible caregivers’ time as above,(32) estimating that 21.5 minutes of preparation time were required, on average, for the breakfast and lunch each day,(33) and that these daily costs would occur for the 20 days of the month that P-EBT was meant to cover (time and travel costs for grocery purchasing were not included, as it was assumed that the P-EBT foods would be purchased during regular household grocery trips).
